# Take-Home Dosing Experiences among Persons Receiving Methadone Maintenance Treatment During COVID-19

**DOI:** 10.1101/2020.08.31.20185249

**Authors:** Mary C. Figgatt, Zach Salazar, Elizabeth Day, Louise Vincent, Nabarun Dasgupta

**Author notes:** Corresponding author: Mary Figgatt, Permanent address: 725 Martin Luther King Blvd, CB# 7505, Chapel Hill, NC 27599.

## Abstract

**Purpose:** Methadone maintenance treatment is a life-saving treatment for people with opioid use disorders (OUD). The coronavirus pandemic (COVID-19) introduces many concerns surrounding access to opioid treatment. In March 2020, the Substance Abuse and Mental Health Services Administration (SAMHSA) issued guidance allowing the expansion of take-home methadone doses. We sought to describe changes to treatment experiences from the perspective of persons receiving methadone at outpatient treatment facilities for OUD.

**Methods:** We conducted an in-person survey among 104 persons receiving methadone from three clinics in central North Carolina in June and July 2020. Surveys collected information on demographic characteristics, methadone treatment history, and experiences with take-home methadone doses in the context of COVID-19 (i.e., before and since March 2020).

**Results:** Before COVID-19, the clinic-level percent of participants receiving any amount of days’ supply of take-home doses at each clinic ranged from 56% to 82%, while it ranged from 78% to 100% since COVID-19. The clinic-level percent of participants receiving a take-homes days’ supply of a week or longer (i.e., ≥6 days) since COVID-19 ranged from 11% to 56%. Among 87 participants who received take-homes since COVID-19, only four reported selling their take-home doses.

**Conclusions:** Our study found variation in experiences of take-home dosing by clinic and little diversion of take-home doses. While SAMSHA guidance should allow expanded access to take-home doses, adoption of these guidelines may vary at the clinic level. The adoption of these policies should be explored further, particularly in the context of benefits to patients seeking OUD treatment.

**Highlights:**

- Methadone take-home dosing of survey participants varied by clinic.
- Diversion of take-home doses was rare.
- Implementation of COVID-19 opioid treatment guidelines should be examined further.

## 1. Introduction

Methadone maintenance treatment is a life-saving treatment for people with opioid use disorder (OUD) (Mattick et al., 2009; Woody et al., 2007). However, the strict requirements of daily in-person, supervised dosing are often burdensome and time-consuming for people participating in treatment (Kourounis et al., 2016). Historically, people receiving methadone are sometimes given take-home doses, however, these doses may be limited to patients with long-term treatment histories and may be provided in doses lasting only one or two days at a time (Walley et al., 2012).

The coronavirus pandemic (COVID-19) has created an urgency for opioid treatment programs (OTPs) to respond by ensuring continued access for existing patients, promoting patient safety, and expanding to new patients (Davis & Samuels, 2020; Del Pozo et al., 2020; Khatri & Perrone, 2020; Krawczyk et al., 2020; Leppla & Gross, 2020). In March 2020, the Substance Abuse and Mental Health Services Administration (SAMHSA) issued guidelines allowing expanded use of take-home doses for more “stable” patients, likely relevant to those who have displayed longer durations of treatment (Substance Abuse and Mental Health Services Administration, 2020).

In an effort to examine changes to methadone take-home policies in the early months of COVID-19, we sought to capture experiences of persons receiving methadone at several methadone clinics in central North Carolina.

## 2. Methods

We conducted a survey among persons receiving methadone prescriptions at methadone clinics in central North Carolina on Monday mornings during June and July 2020. We identified all methadone clinics within a 50-mile radius of Greensboro, North Carolina (n=10) and contacted them in a randomized order. The first three that agreed to participate were included in the study. Two clinics (Clinics A & B) were for-profit and one was non-profit. We estimated needing approximately 100 patients to reach saturation of experiences while balancing COVID-19-related research restrictions. The survey was developed by the authors and reviewed by persons with lived experience of methadone treatment. The survey was also reviewed by clinic staff, but no changes were made upon this review.

On-site recruitment and survey administration was conducted by staff from the North Carolina Survivors Union (NCSU). NCSU is a self-support group of people with lived experience of drug use that operates a drop-in center and harm reduction program in Greensboro, North Carolina. NCSU staff approached patients prior or upon entry to the clinic, and while some patients initially declined, some were verbally encouraged to participate by clinic staff while receiving their dose. Subsequently, several patients chose to participate upon exiting the clinic. Prior to the survey launch, NCSU representatives developed a procedural plan for survey recruitment, including verbal scripts and logistical considerations for on-site recruitment. NCSU representatives approached persons entering the clinic and administered verbal consent to those indicating interest in survey participation. Surveys were administered in the waiting room at Clinic A and outdoors at Clinics B and C. Among the three clinics included in our survey, the percent of persons approached who participated were 79% from Clinic A, 34% from Clinic B, and 54% from Clinic C. Among persons who provided verbal consent, we administered a one-page paper-based questionnaire asking about participants’ demographic characteristics, methadone treatment history, receipt of and experiences with take-home doses, and knowledge of peer’s experiences with take-home doses.

We examined participants’ self-reported experiences before and since COVID-19, which was defined in both the survey and analysis as before and after March 1, 2020. We estimated the duration of treatment at the time of March 1, 2020 by subtracting the time occurring between March 1, 2020 and the date of survey administration from the self-reported duration of the participant’s current treatment episode. We examined frequencies and percentages of participants’ reported specific characteristics. All analyses were performed using SAS 9.4.

This study was reviewed and approved by the University of North Carolina at Chapel Hill Institutional Review Board. People with lived experience were involved in design of the study, questionnaire development, data collection, and interpretation of data.

## 3. Results

### 3.1. Characteristics of Survey Participants

Among 104 survey participants recruited from three methadone clinics in central North Carolina, 54.5% were aged 18-34 years old, 55.9% were men, and 88.2% were non-Hispanic white (Table 1). Approximately 18.1% (n=17) of participants had been receiving methadone in their current treatment episode for less than six months, 27.7% (n=26) had been receiving methadone for six to 12 months, and 54.3% (n=51) had been receiving methadone for more than 12 months. When participants receiving take-home doses were asked about the length of their usual days’ supply, 59.8% (n=52) reported their usual days’ supply was for 1-2 days, 13.8% (n=12) reported 4-5 days, 17.2% (n=15) reported 6-12 days, and 9.2% (n=8) reported more than 12 days.

**Table 1.** Summary of survey self-reported participant demographic characteristics and methadone take-home dosing experiences among participants of three clinics in central North Carolina (N=104).

|  | **N=104** | **%** |
| --- | --- | --- |
| Age |  |  |
| 18-34 | 54 | 54.5% |
| 35-54 | 41 | 41.4% |
| 55+ | 4 | 4.0% |
| Missing | 5 |  |
| Gender |  |  |
| Men | 57 | 55.9% |
| Women | 45 | 44.1% |
| Missing | 2 |  |
| Race/ethnicity |  |  |
| Hispanic/Latino | 1 | 1.0% |
| Non-Hispanic Black | 8 | 7.8% |
| Non-Hispanic white | 90 | 88.2% |
| Other | 3 | 2.9% |
| Missing | 2 |  |
| Duration of methadone treatment in current treatment episode |  |  |
| <6 months | 17 | 18.1% |
| 6 months – 12 months | 26 | 27.7% |
| >12 months | 51 | 54.3% |
| Missing | 10 |  |
| Personally knew of anyone who: |  |  |
| Gave take-home doses to help someone else | 15 | 14.4% |
| Lost or had take-homes doses stolen | 7 | 6.7% |
| Had people they live with access their take-home doses | 5 | 4.8% |
| Skipped take-home doses to save up for later personal use | 11 | 10.6% |
| Hypothetical reasons why people might give away take-home doses: |  |  |
| Saving up for when clinic is closed | 22 | 21.2% |
| Saving up for travel | 30 | 28.8% |
| Helping someone else | 39 | 37.5% |
| Needing money or drugs | 40 | 38.5% |
| Received take-home doses before COVID-19 | 69 | 68.3% |
| Received take-home doses since COVID-19 | 87 | 91.6% |

### 3.2. Reported Take-Home Dosing Experiences

Among all participants, 68.3% (n=69) had received take-home doses at any point prior to the beginning of COVID-19-related responses (i.e., before March 2020). This proportion increased to 91.6% (n=87) receiving take-home doses since the widespread onset of COVID-19. Only six (6.9%) of the 87 participants receiving take-home doses since COVID-19 reported either selling or sharing their doses. Specifically, four persons (4.6%) reported selling their doses and three (3.4%) reported sharing their doses. Among participants receiving take-homes doses, 71.3% (n=62) reported storing doses in a child-resistant or locked container, 64.4% (n=56) reported having a safe storage location at home, while only 3.4% (n=3) reported that other people residing at their home could access their doses. Among all participants, regardless of whether they received take-home doses, 14.4% (n=15) reported knowing someone who gave away doses to help someone else. Only five (4.8%) participants reported knowing someone who had other people they lived with get into their doses. The most reported hypothetical reasons for giving away doses included: needing money or drugs (38.5%), helping someone else like a friend (37.5%), and saving up for travel (28.8%).

### 3.3. Variation in Take-Home Dosing by Clinic and Treatment Duration

The percent of participants receiving take-home doses before COVID-19 ranged from 56.1% in Clinic C to 82.1% in Clinic A (Figure 1a). Participants receiving any take-home doses since COVID-19 varied from 92.6% in Clinic A, 100% in Clinic B, and 78.0% in Clinic C. Participants receiving take-homes of a week supply or longer (six or more days) since COVID-19 varied from 55.6% in Clinic A, 13.3% in Clinic B, and 10.5% in Clinic C. While two-thirds of participants with treatment durations longer than 12 months received take-homes prior to COVID-19, 56.3% of participants with treatment duration less than six months received any amount of take-homes prior to COVID-19 (Figure 1b). When examining the estimated duration of treatment at the beginning of the widespread COVID-19 response in the U.S. (i.e. March 1, 2020), the receipt of any amount of take-homes since COVID-19 was 90.0% (n=9) among those with an estimated treatment duration of less than 6 months at this time, 94.4% (n=17) among those with 6 to 12 months of treatment, and 91.5% (n=43) among those with longer than 12 months of treatment.**4**.

**Figure 1.**
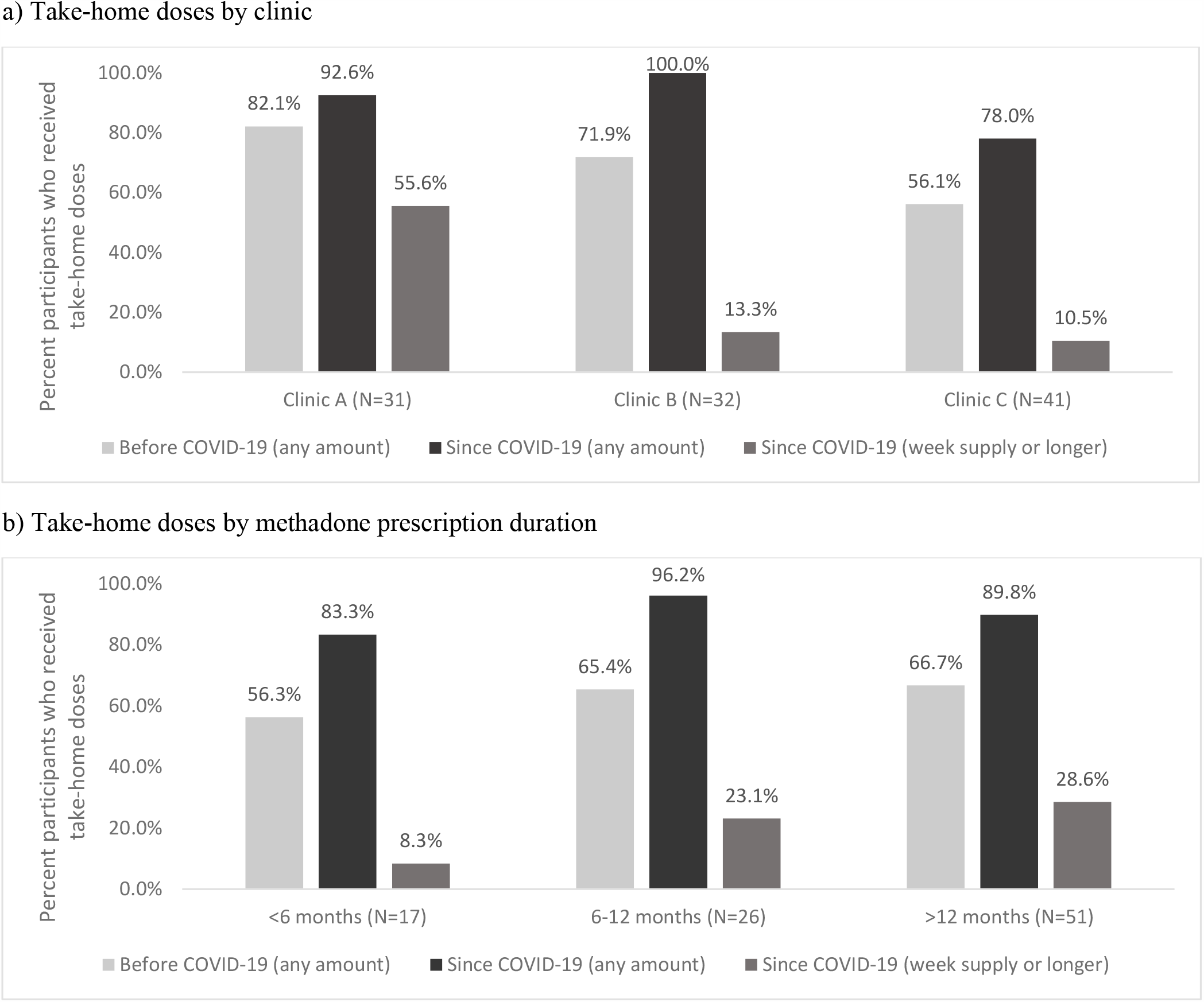
Percent of survey participants take-home dose experiences pre- and post-COVID-19 by clinic and treatment duration. Before COVID-19 was defined as prior to March 2020. Since COVID-19 was defined as since March 2020. Length of take-home doses was defined by asking participants the typical days’ supply of take-home doses they received since March 2020. Duration of treatment was identified by asking participants about the length of methadone treatment in their current treatment episode. In Figure 1a, data were unavailable for: 4 of the 31 participants at Clinic A, 2 of the 32 participants at Clinic B, and 3 of the 41 participants at Clinic C. In Figure 1b, data were unavailable for: 5 of the 17 participants in the <6 month category for take-homes since COVID-19, 0 of the 26 participants in the 6-12 month category for take-homes since COVID-19, and 2 of the 51 participants in the >12 month category.

## Discussion

Our study found variation in the proportion of persons receiving take-home doses since COVID-19 at several clinics in central North Carolina. While our study is specific to methadone maintenance treatment in three clinics, these results may suggest a need for a better understanding of how the SAMHSA directive was implemented during COVID-19. The COVID-19 pandemic creates substantial urgency in improving and expanding access to persons seeking OUD treatment (Davis & Samuels, 2020; Peavy et al., 2020). In addition, COVID-19 presents an opportunity to re-examine dated regulations surrounding OUD treatment (del Pozo & Beletsky, 2020; Krawczyk et al., 2020).

Given the SAMSHA guidelines allowing expanded access to take-home doses (Substance Abuse and Mental Health Services Administration, 2020), we expected to identify less variation in take-home practices by clinic. In fact, some survey participants reported preferring when the clinic provided take-home doses every other day, indicating that some clinics may have provided take-home doses at one point but stopped by the time the survey was administered in June and July 2020. Very few participants reported selling or giving away their take-home doses. Conversations surrounding diversion have historically focused on persons selling their methadone doses (Institute of Medicine (US) Committee on Federal Regulation of Methadone Treatment, 1995). However, our survey suggests selling doses may be relatively uncommon. In addition, reasons for giving away or otherwise not taking methadone take-homes as prescribed may follow a much more innocuous narrative than diversion for profit, such as saving up doses for travel or helping someone else. A continuous focus on diversion as a central rationale for restricting take-home dosing may increase stigma and further marginalize people who are prescribed methadone for OUD treatment. Instead, our results suggest the need to examine the benefits of receiving take-home doses on treatment, recovery, and general well-being of person receiving methadone.

Our study has several limitations. First, due to our study being administered on a single day per clinic, we did not capture true prevalence of take-home doses overall among survey participants. Rather, these results can be used to assess variation in persons receiving take-home doses by clinic and treatment duration. Second, participation in the survey varied by clinic. We believe this was due to some participants being more strongly encouraged to participate by clinic staff and possibly the location of survey administration. For example, surveys were administered while participants waited to be seen by clinic staff at Clinic A, whereas surveys were administered to participants as they entered and exited the clinic at Clinics B and C. Finally, it is possible social desirability bias was present in the self-report of sensitive questions, such as those related to diversion or safe storage procedures. Very low (7%) endorsement of diversion in this survey challenges the loss-prevention orientation of methadone programs. Yet, given the in-clinic milieu of questionnaire deployment and concerns regarding repercussions leading to disruptions in treatment, we cannot preclude self-report bias or that respondents who chose to participate may have self-selected for higher adherence behaviors. Clinic staff were not able to see individual survey results, possibly mitigating the former. However, our survey was administered by a separate organization (NCSU) comprised of persons with lived experience of drug use and community advocates, which may result in participants being be more comfortable disclosing sensitive behaviors to this group rather than clinic staff.

## 5. Conclusions

Our findings have implications for evaluations of the SAMHSA pandemic directive. We observed considerable heterogeneity in take-home practices between clinics, suggesting differences in interpretation. Additional research is needed to explore the extent and possible reasons of this heterogeneity in greater detail. Future studies should also consider exploring clinic administrator perspectives and behaviors. OTPs should reduce barriers to treatment during COVID-19 and consider expanding access to take-home doses while providing harm reduction messaging to their patients. Instituting additional barriers to treatment may come at the cost of lives; therefore, we should make treatment more accessible, not more restrictive.

## Data Availability

Data are not currently available for external use. However, if there is interest in obtaining additional information, please contact one of the authors.

## Acknowledgements

We are appreciative of Elizabeth Joniak-Grant, Maryalice Nocera, and Toska Cooper for their support of this work.

